# Enhancing MR imaging driven Alzheimer’s disease classification performance using generative adversarial learning

**DOI:** 10.1101/2020.07.22.20159814

**Authors:** Xiao Zhou, Shangran Qiu, Prajakta S. Joshi, Chonghua Xue, Ronald J. Killiany, Asim Mian, Sang P. Chin, Rhoda Au, Vijaya B. Kolachalama, for the Alzheimer’s Disease Neuroimaging Initiative

**Affiliations:** Section of Computational Biomedicine, Department of Medicine, Boston University School of Medicine, Boston, MA, USA; Department of Computer Science, College of Arts & Sciences, Boston University, MA, USA; Department of Physics, College of Arts & Sciences, Boston University, MA, USA; Department of Anatomy and Neurobiology, Boston University School of Medicine, Boston, MA, USA; Department of General Dentistry, Boston University School of Dental Medicine, Boston, MA, USA; Department of Radiology, Boston University School of Medicine, Boston, MA, USA; Department of Neurology, Boston University School of Medicine, Boston, MA, USA; Boston University Alzheimer’s Disease Center, Boston, MA, USA; Department of Brain and Cognitive Science, Massachusetts Institute of Technology, Cambridge, MA, USA; Center of Mathematical Sciences & Applications, Harvard University, Cambridge, MA, USA; The Framingham Heart Study, Boston University School of Medicine, Boston, MA, USA; Department of Epidemiology, Boston University School of Public Health, Boston, MA, USA; Faculty of Computing & Data Sciences, Boston University, Boston, MA, USA

**Keywords:** Magnetic resonance imaging, Alzheimer’s disease, Generative adversarial network, Fully convolutional network

## Abstract

**Background:** Generative adversarial networks (GAN) can generate images of improved quality but their ability to augment image-based classification tasks is not fully explored.

**Purpose:** We evaluated if a modified GAN can learn from MRI scans of multiple magnetic field strength to enhance Alzheimer’s disease (AD) classification performance.

**Materials and methods:** T1-weighted brain MRI scans from 151 participants of the Alzheimer’s Disease Neuroimaging Initiative (ADNI), who underwent both 1.5 Tesla (1.5T) and 3 Tesla imaging at the same time were selected to construct a GAN model. This model was trained along with a three-dimensional fully convolutional network (FCN) using the generated images (1.5T*) as inputs to predict AD status. Quality of the generated images was evaluated using signal to noise ratio (SNR), Blind/Referenceless Image Spatial Quality Evaluator (BRISQUE) and Natural Image Quality Evaluator (NIQE). Data from the Australian Imaging, Biomarker & Lifestyle Flagship Study of Ageing (AIBL, n=107), and the National Alzheimer’s Coordinating Center (NACC, n=565) was used for model validation.

**Results:** The mean quality of the generated (1.5T*) images was consistently higher than the 1.5T images, as measured using SNR, BRISQUE and NIQE on the validation datasets. The 1.5T*-based FCN classifier performed better than the FCN model trained using the 1.5T scans.

Specifically, the mean area under curve increased from 0.907 to 0.932, from 0.934 to 0.940 and from 0.870 to 0.907 on the ADNI test, AIBL and NACC datasets, respectively.

**Conclusion:** This study demonstrates that GAN frameworks can be constructed to simultaneously improve image quality and augment classification performance.

**Key points:**

- Our proposed generative adversarial network used 1.5 and 3 Tesla MR scans of the brain to generate images of improved quality, as estimated using no-reference image quality algorithms.
- Classification models of Alzheimer’s disease risk developed using the generated images had higher classification performance than the models developed using the original 1.5 Tesla scans.

## Introduction

Rapid improvements in neuroimaging techniques such as magnetic resonance imaging (MRI) have led to more sensitive method of identifying neurodegeneration associated with Alzheimer’s disease (AD) pathology before the onset is irreversible (1). Early evaluation of the pathophysiological changes on MR images could potentially facilitate the creation of new models of disease progression, the discovery of new treatments, and help patients, families, and clinicians prepare for the later stages of the disease. The pace of research, however, is not determined only by the rate at which a new imaging technology is developed. AD is slowly progressive and because early stages of the disease are clinically asymptomatic, image-based screenings of at-risk individuals are usually carried out after neurodegeneration has begun. This means that the ongoing national longitudinal studies such as Alzheimer’s Disease Neuroimaging Initiative (ADNI) (2), Australian Imaging, Biomarker & Lifestyle Flagship Study of Ageing (AIBL) (3), and the National Alzheimer’s Coordinating Center (NACC) (4), dedicated to track AD progression from its clinically pre-symptomatic phase forward can advance no faster than the disease itself. Additionally, these studies make use of increasingly advanced imaging technology as it becomes available, rendering it difficult to distinguish biological changes due to disease from brain scans that are unchanged but have improved image quality. Thus, even though longitudinal assessment is essential for both diagnosing AD and understanding its mechanisms, the acquisition of images at different time points with a different technology poses inherent technical bias. AD research requires a technique that would enable comparison of longitudinal sequences of images that conform to a common technological standard of precision. Additionally, there is a need for statistically significant number of baseline image sequences prior to the onset of disease that can be analyzed without having to wait for the neuropathological features of the disease to develop to an irreversible stage.

One possible solution to partially addressing this issue is using generative adversarial learning (5), which is an emerging technique in machine learning that incorporates a system of two neural networks that compete with each other in a zero-sum game framework. Since its introduction, this framework has shown to improve the quality of imaging data in general, and medical images in particular (6). Here, we evaluated if a generative adversarial network (GAN) can be developed to generate images of improved quality and also augment performance of a classifier trained using the generated images. To achieve this goal, we processed brain MRI scans of multiple magnetic field strengths (1.5 Tesla (1.5T) and 3 Tesla (3T)) from the ADNI dataset, and also obtained access to 1.5T MRI scans from the AIBL and NACC datasets. Using these data, we addressed the following objectives. First, the deep learning framework needs to generate images that have improved quality. To achieve this, we trained a GAN model using 1.5T and 3T scans obtained around the same time on the same set of individuals. We then used well-known image quality metrics to compare the original scans and the generated images.

Second, the deep learning framework needs to more accurately predict the class label than what one could achieve using the original scans. To achieve this, we used the generated images to construct a fully convolutional network (FCN) that discriminated between cases who had AD from those who had normal cognition. For comparison, we also generated an independent FCN model using the original 1.5T scans to predict AD status. Validation of the FCN models was performed using data from the AIBL and NACC studies.

## Materials and methods

### Study population and MRI scan parameters

We obtained access to T1-weighted MRI scans from the ADNI (n=417), AIBL (n=107) and NACC (n=565) cohorts (Table 1). For a subset of the ADNI data (n=151), both 1.5 Tesla (1.5T) and 3 Tesla (3T) scans taken at the same time were available, and 1.5T scans were available for the other cohorts. All the MRI scans considered for this study were performed on individuals within ±6 months from the date of clinical assessment.

**Table 1:** Study population and characteristics. Three independent datasets including (a) the Alzheimer’s Disease Neuroimaging Initiative (ADNI) dataset, (b) the Australian Imaging, Biomarker & Lifestyle Flagship Study of Ageing (AIBL), and (c) the National Alzheimer’s Coordinating Center (NACC)) were used for this study.

|  | ADNI 1.5T |  |  | ADNI 3T |  |  | NACC |  | AIBL |  |
| --- | --- | --- | --- | --- | --- | --- | --- | --- | --- | --- |
| **Diagnosis** | **NC** | **MCI** | **AD** | **NC** | **MCI** | **AD** | **NC** | **AD** | **NC** | **AD** |
| **Number of cases** | 229 | 69 | 188 | 47 | 69 | 35 | 356 | 209 | 93 | 14 |
| **Age (median + range)** | 76<br>[60, 90] | 76<br>[55, 88] | 76<br>[55, 91] | 75<br>[70-86] | 76<br>[55, 88] | 72<br>[57, 89] | 74<br>[56, 94] | 77<br>[55, 95] | 71<br>[61, 86] | 73<br>[58, 82] |
| **Gender, male (percent age)** | 119<br>(51.9 6%) | 39<br>(56.5 2%) | 101<br>(53.7 2%) | 18<br>(38.2 9%) | 39<br>(56.5 2%) | 12<br>(34.2 9%) | 126<br>(35.3 9%) | 95<br>(45.4 5%) | 48<br>(51.6 1%) | 6<br>(42.8 6%) |
| **Education (median + range)** | 16<br>[6, 20] | 16<br>[6, 20] | 16<br>[4, 20] | 16<br>[7, 20] | 16<br>[6, 20] | 14<br>[7, 20] | 16<br>[0, 22] | 14.5<br>[2, 24] | N.A. | N.A. |
| **APOE+ (percent age)** | 61<br>(26.6 4%) | 33<br>(47.8 3%) | 124<br>(65.9 6%) | 13<br>(27.6 6%) | 33<br>(47.8 3%) | 24<br>(68.7 5%) | 102<br>(28.6 5%) | 112<br>(53.5 9%) | 1<br>(1.01 %) | 1<br>(7.17 %) |
| **MMSE (median + range)** | 29<br>[25, 30] | 26<br>[24, 30] | 23.5<br>[18, 28] | 30<br>[26, 30] | 26<br>[24, 30] | 23<br>[20, 27] | 29<br>[20, 30] | 22<br>[0, 30] | 29<br>[25, 30] | 18<br>[6, 22] |

ADNI is a longitudinal multicenter study designed to develop clinical, imaging, genetic, and biochemical biomarkers for the early detection and tracking of AD (7). AIBL, launched in 2006, is the largest study of its kind in Australia and aims to discover biomarkers, cognitive characteristics, and lifestyle factors that influence the development of symptomatic AD (3).

Finally, NACC, established in 1999, maintains a large relational database of standardized clinical and neuropathological research data collected from AD centers across the US (8).

The MRI scans used in this study from the ADNI dataset are from the baseline visit. For a subset of the ADNI participants (n=151), both 1.5T and 3T scans taken at the same study visit were available. Scanning on ADNI focused on consistent longitudinal structural imaging on 1.5T scanners using T1-weighted sequences, and a group of subjects were also scanned using the same protocol on 3T scanners. For each scanning sequence (MP-RAGE), the geometry defining the field of view at reconstructed resolution was 208×240×256 mm at 1×1×1 mm. The timing parameters included TE=min full echo, TR=2300, T1=900 and the approximate runtime was 6.2 minutes. These scans from the ADNI data were used for GAN model training and testing. The remaining participants from the ADNI, AIBL and NACC studies had 1.5T MRI scans available and were used for FCN model testing. Note that the AIBL study also used an imaging protocol similar to the ADNI study. However, for the NACC study, a single protocol was not available as it is a collection of scans from several AD centers.

### Image registration and data normalization

We used the linear registration tool from the FSL package (University of Oxford, UK) to register raw MRIs based on the MNI152 template (ICBM 2009c Nonlinear Symmetric template, McGill University, Canada). After registration, we applied z-score normalization on all the voxels of the whole brain volume and cleared the background noise. A depth-first search algorithm was implemented to expand the search area from initial corner locations into inner regions until skull was encountered. An intensity threshold was set and the search regions could only expand to voxels with intensity lower than the threshold. As a result, the signal of the sub-cortical fat which is higher than the threshold prohibited the search region leaking into the brain. We then set the background noise value to ‘−1’, which is the averaged background intensity value. Lastly, to eliminate outlier voxels with high intensity, we clipped every voxel to the range: [−1, 2.5], by setting any voxel with intensity lower than ‘−1’ to value ‘−1’, and any voxel with intensity higher than ‘2.5’ to ‘2.5’.

### Deep learning framework

Original 1.5T scans from the ADNI data on individuals who received both a 1.5T and 3T scan were fed into the generator that created images, and the discriminator was used to compare the generated images (1.5T*) with the original 3T images. At the same time, a fully convolutional network (FCN) was trained using the generated 1.5T* images to discriminate AD cases with the ones who have normal cognition (NC). The goal was to find a mapping function which results in the creation of an image mask, which when added to a 1.5T scan generates an image (1.5T*) that has the same or better quality, and leads to a more accurate prediction of AD status. While the generator is using the mapping function to create a mask, the discriminator is attempting to distinguish between the generated 1.5T* image and the original 3T image in an adversarial fashion. Concomitantly, the FCN model is attempting to distinguish between AD and NC cases. In essence, both the GAN and the FCN model training is performed simultaneously while minimizing the GAN and classifier losses (Figure 1). This was achieved by allowing the gradient calculated from the FCN classification loss to propagate back to the generator to implicitly convey the disease related information to the generator. The expectation is that the classification loss that is propagated provides a momentum for the generator to generate images that contributed to lower cross-entropy loss and thus facilitated better image classification.

**Figure 1:**
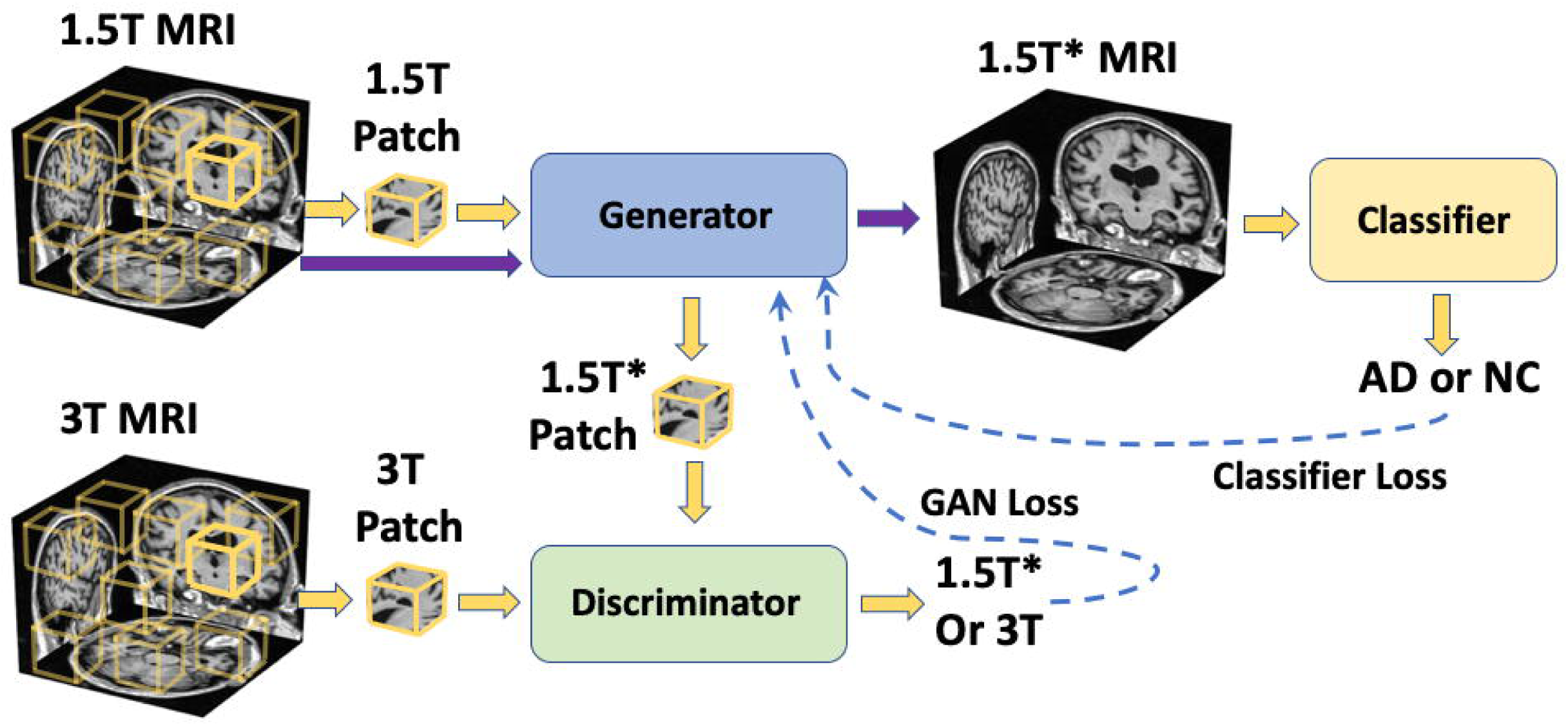
Schematic of the overall deep learning strategy. The generative adversarial network (GAN) uses 1.5T and 3T scans of the same individual taken at the same time to generate images (1.5T*). The fully convolutional network (FCN) model uses the 1.5T* images to predict Alzheimer’s disease (AD) status. Both the GAN and FCN models were trained simultaneously by backpropagating GAN and the classifier losses.

The generator of the GAN model consists of three 3D convolutional blocks in which each convolutional operation was followed by batch normalization and rectified linear unit (ReLu) activation. In each convolution layer, the stride and kernel size were set at 1 and 3, respectively, and padding as 2, 0, and 1, to guarantee the output from the generator to have the same size as input so that we could directly add the mask on the 1.5T scan. The discriminator of the GAN model is fully convolutional, consisted of 5 convolutional blocks in which 3D convolution operations were followed by batch normalization and LeakyReLu activation. The model was trained with losses from the discriminator and the FCN classifier, as well as an additional L_1_-norm loss calculated between the original 1.5T image and the generated image. More details are described in the supplement.

The FCN model was trained to predict AD status using the generated images (1.5T*) as inputs. For comparison, we trained another FCN model using 1.5T scans of the same individuals to predict AD status. The NINCDS-ADRDA criterion was used to define the AD status (9). The first FCN model was constructed using the 1.5T scans on the entire ADNI dataset (n=417), and the second FCN model used the 1.5T* images and was trained together with the GAN model. For each FCN model, the cases were randomly divided into three groups in the ratio of 3:1:1 for training, validation and testing, respectively.

We used patch-wise training for both the GAN and the FCN models. The training process using this strategy was less computationally intensive and allowed us to use neural networks with larger capacity given a total memory budget. More specifically, we randomly sampled patches of size 47×47×47 from the whole volume as inputs to the deep learning framework. Patches from the 1.5T scans were sent into the generator, and the discriminator then attempted to differentiate between the 1.5T* patch and 3T patches. The FCN model then used the 1.5T* patches as input to predict AD status. With the strategy of randomly sampling patches over the whole volume, a degree of data augmentation was achieved because the model was trained with more variance of the inputs sampled from various locations. Similar FCN frameworks have been used recently to generate high performance AD classification models (10).

### Data partitioning

The GAN model was constructed on a subgroup of ADNI data (n=151), which contained both 1.5T and 3T scans from same individuals taken at the same time. This subgroup of the ADNI data was randomly split into training, validation and testing in the ratio of 3:1:1. The GAN model was trained on the training part of the 151 cases, and the model was saved at the time point where the image quality metric SNR on the validation set was the highest. The saved GAN model was subsequently evaluated on the test data by generating 1.5T* MRI for those cases.

For the FCN model, the cases from the ADNI dataset were randomly divided into three groups in the ratio of 3:1:1 for training, validation and testing, respectively. The validation part was used to identify and save the model at the optimal epoch; and the testing split of the data was used to evaluate the model performance on the data which it had never seen. Additionally, we evaluated our models on 2 external datasets (AIBL & NACC). Note that all the 151 cases used for GAN training, validation and testing were also used for FCN training in order to blind the GAN model or the FCN models to the validation and testing data.

### Quality metrics

We used signal to noise ratio (SNR) as well as no-reference algorithms including Blind/Referenceless Image Spatial Quality Evaluator (BRISQUE) (11), and Natural Image Quality Evaluator (NIQE) (12), to compare the differences between the original scans and generated images.

For SNR, we computed the average values of pixel intensity and divided it by its standard deviation. BRISQUE focuses on quantifying spatial distortion, such as ringing, blur or blocking, from natural images. Certain regular statistical properties of natural images could be influenced by the presence of distortions. The BRISQUE evaluator was developed by learning the difference between original natural images and distorted images using the LIVE IQA database (13, 14). NIQE is also a non-reference evaluator, which quantifies image quality according to the level of distortions. The difference of NIQE compared with BRISQUE is that NIQE does not require distorted images as a prior, thus could learn only from undistorted images. Lower BRISQUE and NIQE scores indicate better image quality.

### Performance metrics for classification

We generated sensitivity-specificity (SS) and precision-recall (PR) curves based on model predictions on the ADNI test data as well as on the other independent datasets (AIBL and NACC). For each SS and PR curve, we also computed the area under curve (AUC) values. Additionally, we computed sensitivity, specificity and F1-score on each set of model predictions. Here, TP denotes true positive values, and FP and FN denote false-positive and false-negative cases, respectively. Both the 1.5T- and 1.5T*-based classification models were trained 25 times with various random seeds and 95% confidence intervals were generated.

### Statistical analysis

A series of analyses were performed to evaluate the image quality and to assess the AD classification performance. To evaluate the mean difference in image quality generated by the GAN model, we performed analysis of variance (ANOVA) on the ADNI test data. Image quality was assessed using SNR, BRISQUE and NIQE. Specific group differences between 1.5T, 3T and 1.5T* images were evaluated using the post hoc Tukey test (Supplement Table 1). The effect of age, education, gender, MMSE scores, ApoE4 status and type of scanner on SNR, BRISQUE and NIQE were evaluated using a stepwise forward selection process using the ‘GLMSELECT’ function (SAS Software) followed by analysis of covariance (ANCOVA). Lastly, we used the t-test to assess whether the mean image quality of the 1.5T group was different from the 1.5T* group in the NACC and AIBL datasets.

## Results

Volumetric patch-level training on the 1.5T and 3T scans allowed training of volumetric masks, which then resulted in the generation of 1.5T* volumetric images. For ease of visualization, we selected the center slice of a single subject and retrieved a two-dimensional mask from the learned volume and compared both the original and generated images (Figure 2). This figure demonstrates that captured differences between the 1.5T and 1.5T* images were subtle and distributed throughout the region. When metrics such as SNR were used, there was a 9.6% improvement (1.31 to 1.45), on the mean image quality on the ADNI test data between the 1.5T and 1.5T* images. We also found that mean SNR improved by about 11.1% (1.26 to 1.40) and 11.7% (1.28 to 1.43) on the AIBL and NACC data, respectively.

**Figure 2:**
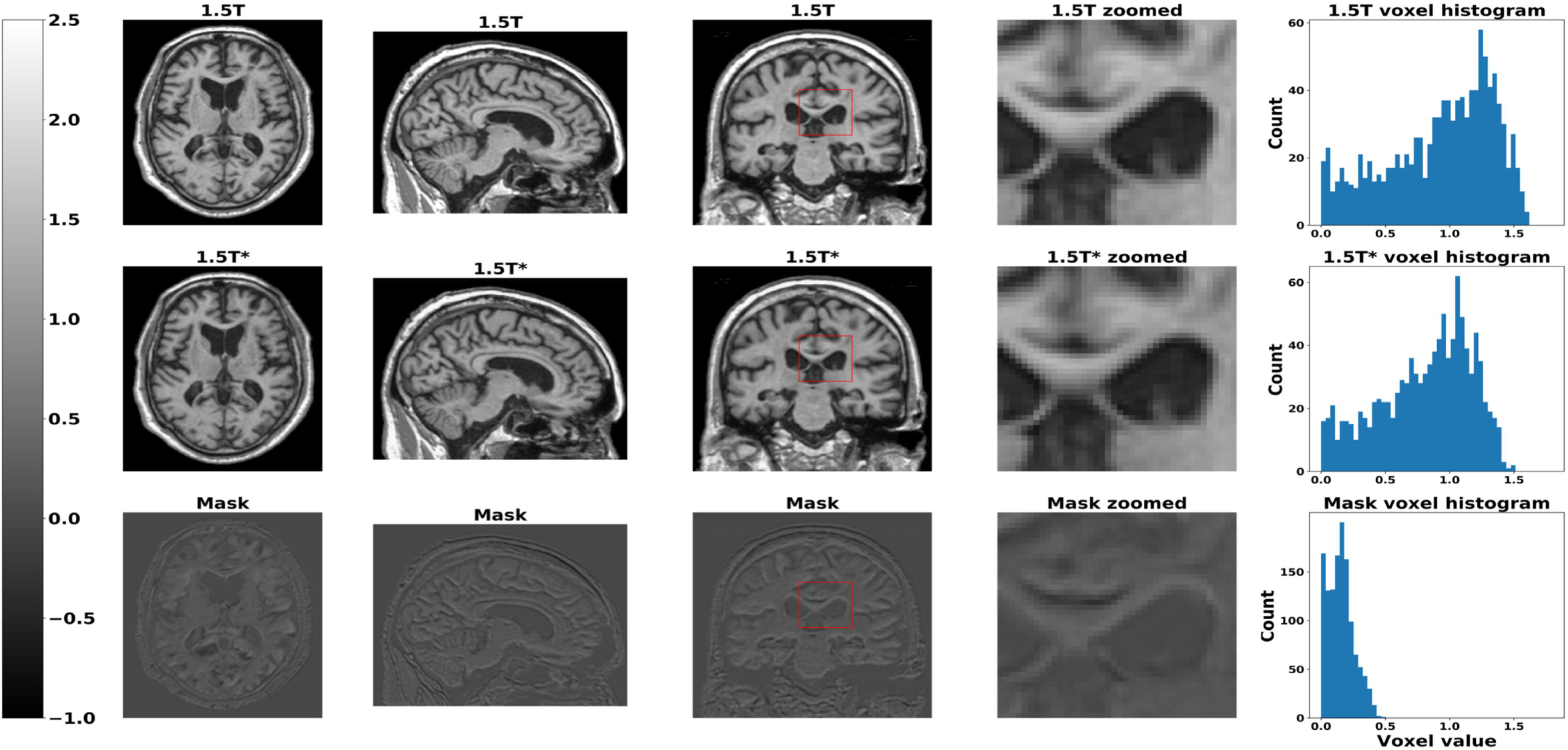
Original and generated images and corresponding masks. Axial, sagittal and coronal views of a single subject are shown in the first, second and third columns, respectively. The first row corresponds to the original 1.5T slices, the second row corresponds to the generated images (1.5T*) and the third row corresponds to the difference between 1.5T and 1.5T* images, denoted as the mask. We also showed the same zoomed-in region from 1.5T, 1.5T* images and the mask in the fourth column to reveal the difference between 1.5T and 1.5T* images. Additionally, we presented histograms of the voxel values within the zoomed-in region in the fifth column of the plot.

Absolute measures of image quality using perceptual quality metrics provided more insight on the differences between 1.5T and 1.5T* images. When BRISQUE metric was used, there was 8.3% improvement in the mean image quality on the ADNI test data (49.06 to 44.97), about 10.0% improvement in the mean image quality on the AIBL data (44.91 to 40.44), and 9.0% mean image quality improvement on the NACC data (47.79 to 43.48). Note that lower BRISQUE score indicates better quality. When NIQE score was used to compare images, there was 16.8% improvement in the mean image quality on the ADNI test data (7.33 to 6.1), 14.3% improvement in the mean image quality on the AIBL data (7.21 to 6.18), and 16.3% mean image quality improvement on the NACC data (7.99 to 6.69). Similar to the BRISQUE metric, lower NIQE score indicates better quality. These no-reference metrics provided an objective way to evaluate the quality of the generated images on different cohorts (AIBL & NACC), which then grounded our hypothesis that the GAN model can learn from images of multiple magnetic field strength to improve image quality.

In order to determine if there was an overall difference in the mean quality of images produced by the GAN model, we performed ANOVA for the SNR, BRISQUE and NIQE metrics computed on the ADNI test data (Figure 3, Supplement Figure 1). We found a significant overall difference in image quality between the 1.5T, 3T and 1.5T* groups using SNR (F=229.66, p<0.0001), BRISQUE (F=10.80, p<0.0001) and NIQE (F=27.95, p<0.0001). To identify the between-group differences for the SNR, BRISQUE and NIQE metrics, we used the Tukey’s post hoc procedure (Supplement Table 1). We found that the 1.5T* group had significantly better image quality compared to 1.5T scans across all three image quality metrics, SNR (p<.0001), BRISQUE (p<.0001), and NIQE (p<.0001). On the other hand, the mean image quality in the 1.5T* category was significantly better than the 3T scans on the SNR metric (p<.0001), but not on the BRISQUE (p=0.1074) and NIQE (p=0.82) metrics.

**Figure 3:**
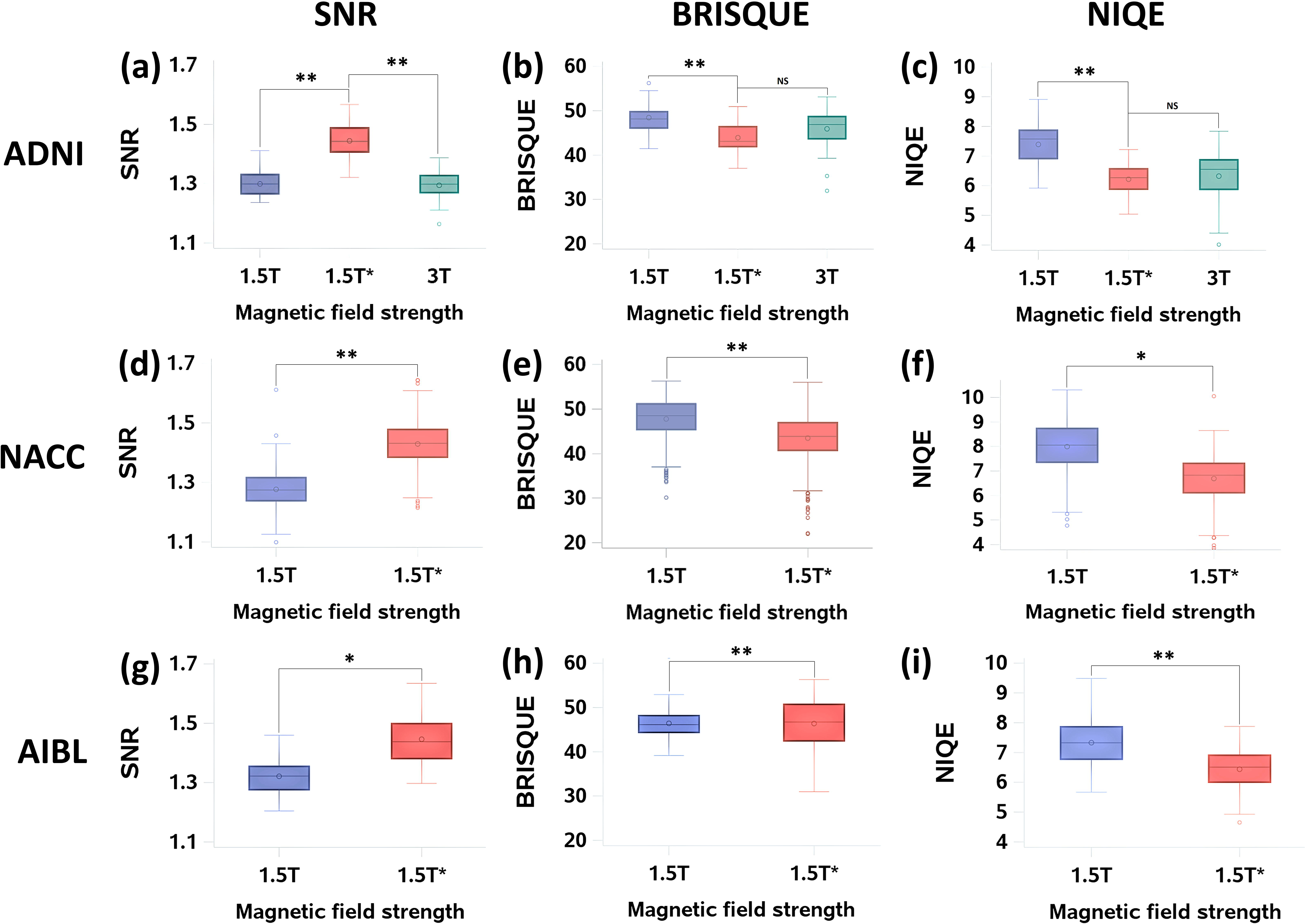
Image quality analysis. Metrics such as SNR as well as no-reference algorithms including BRISQUE and NIQE were used to evaluate the quality of the generated images (1.5T*) and compare them with the quality of the original scans (1.5T and 3T). The metrics were computed independently on the MRI scans from each study cohort (ADNI-test (a-c), NACC (d-f), AIBL (g-i)). Lower value of the metrics indicates improved quality. The symbol ‘*’ indicates p<0.001 and ‘**’ indicates p<0.0001.

A stepwise forward selection process using the ‘GLMSELECT’ function (SAS Software) was used to evaluate the effect of age, education, gender, MMSE scores, ApoE4 status and type of scanner on SNR, BRISQUE and NIQE. For SNR, ‘age’ was statistically significant (p=0.0003). For BRISQUE, ‘years of education’ (p=0.04) and ‘scanner type’ (p=0.003) were statistically significant, and for NIQE, ‘age’ (p=0.001), ‘years of education’ (p=0.01) and ‘scanner type’ (p=0.006) were statistically significant. Lastly, an analysis of covariance was performed by adjusting for the above-mentioned covariates. The mean difference in image quality assessed by SNR in both the 1.5T and 3T groups was 0.15 units lower than the 1.5T* scans (p<.0001) after adjusting for age. On adjusting for years of education, the mean difference in image quality between 1.5T and 1.5T* images assessed by the BRISQUE metric was 4.53 (p<.0001) and between 1.5T* and 3T images was 2.00 (p=0.04). Lastly, after adjusting for age, years of education and type of scanner, the mean difference in image quality assessed by NIQE between 1.5T and 1.5T* images was 1.16 units (p<.0001), however the association was not significant between 1.5T* and 3T images (p=0.54).

The generated images also led to consistent, high AD classification performance across the external datasets, at least as demonstrated by area under the sensitivity-specificity and the precision-recall curves (Figure 4a & Figure 4b). The 1.5T*-based FCN model demonstrated improved performance on both the AIBL and NACC datasets, using most of the computed performance metrics (Table 2a & Table 2b). It is worth noting that of Matthews correlation coefficient (MCC) values, which is generally regarded as a balanced measure which can be used even if the classes are of very different sizes, the mean MCC value increased by 19.6% on the AIBL dataset (0.5757 to 0.6884), and 9.2% on the NACC dataset (0.6032 to 0.6585), respectively. As for F1-score, which is generally used as a weighted score of the model’s performance, also increased by 18.8% on the AIBL dataset (0.6126 to 0.7276). The 95% confidence intervals for the FCN models show that the model predictions were fairly consistent across different runs and different between the 1.5T- and 1.5T*-based FCN models (Table 3).

**Table 2:** Performance of the FCN models. Accuracy, sensitivity, specificity, F1-score, and Matthew’s correlation coefficient are computed for the FCN models that used (a) 1.5T scans and (b) 1.5T* images, respectively.

| **1.5T** | **Accuracy** | **Sensitivity** | **Specificity** | **F-1** | **MCC** |
| --- | --- | --- | --- | --- | --- |
| **ADNI test** | 0.8398±0.0238 | 0.7363±0.0514 | 0.9209±0.0385 | 0.7972±0.0325 | 0.6766±0.0492 |
| **AIBL** | 0.8873±0.0647 | 0.6309±0.1393 | 0.9259±0.0874 | 0.6126±0.1016 | 0.5757±0.1086 |
| **NACC** | 0.8157±0.0224 | 0.6739±0.0605 | 0.8989±0.0616 | 0.7301±0.0221 | 0.6032±0.0425 |

(b)
| **1.5T\*** | **Accuracy** | **Sensitivity** | **Specificity** | **F-1** | **MCC** |
| --- | --- | --- | --- | --- | --- |
| **ADNI test** | 0.8210±0.0143 | 0.7411±0.0312 | 0.8895±0.0114 | 0.7923±0.0195 | 0.6416±0.0280 |
| **AIBL** | 0.9293±0.0132 | 0.7143±0.0000 | 0.9617±0.0152 | 0.7276±0.0365 | 0.6884±0.0449 |
| **NACC** | 0.8429±0.0069 | 0.7393±0.0180 | 0.9037±0.0134 | 0.7768±0.0096 | 0.6585±0.0149 |

**Table 3:** Confidence intervals of model performance. (a) 95% confidence intervals of the SS curves for 1.5T-based model, 1.5T*-based model and difference of AUCs between 1.5T* and 1.5T-based models. (b) 95% confidence intervals of PR curves for 1.5T-based model, 1.5T*-based model and difference of AUCs between 1.5T* and 1.5T-based models.

|  | **1.5T** | **1.5T\*** | **1.5T\* - 1.5T** |
| --- | --- | --- | --- |
| **ADNI test** | [0.8968, 0.9172] | [0.9304, 0.9336] | [0.0203, 0.0297] |
| **AIBL** | [0.9258, 0.9422] | [0.9373, 0.9427] | [0.0021, 0.0097] |
| **NACC** | [0.8590, 0.8810] | [0.9058, 0.9082] | [0.0314, 0.0412] |

(b)
|  | **1.5T** | **1.5T\*** | **1.5T\* - 1.5T** |
| --- | --- | --- | --- |
| **ADNI test** | [0.9022, 0.9218] | [0.9324, 0.9356] | [0.0175, 0.0265] |
| **AIBL** | [0.7347, 0.7833] | [0.7467, 0.7593] | [-0.0170, 0.0051] |
| **NACC** | [0.8268, 0.8512] | [0.8768, 0.8792] | [0.0334, 0.0443] |

**Figure 4:**
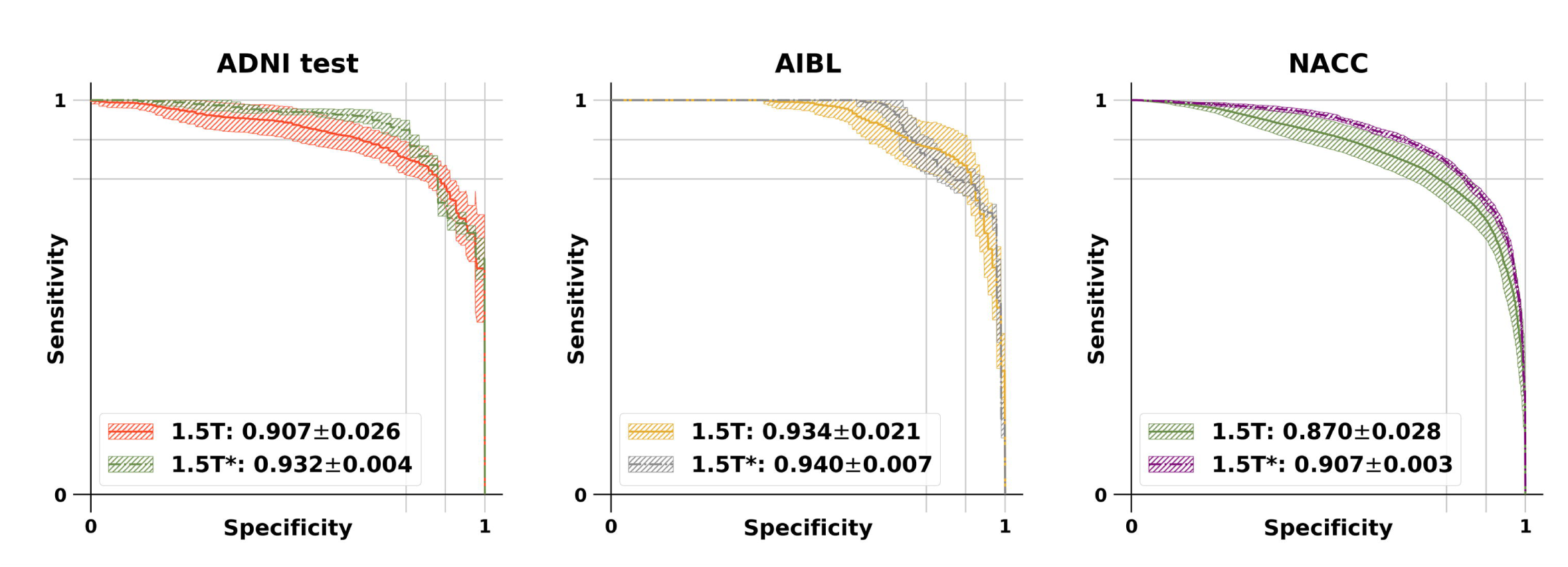
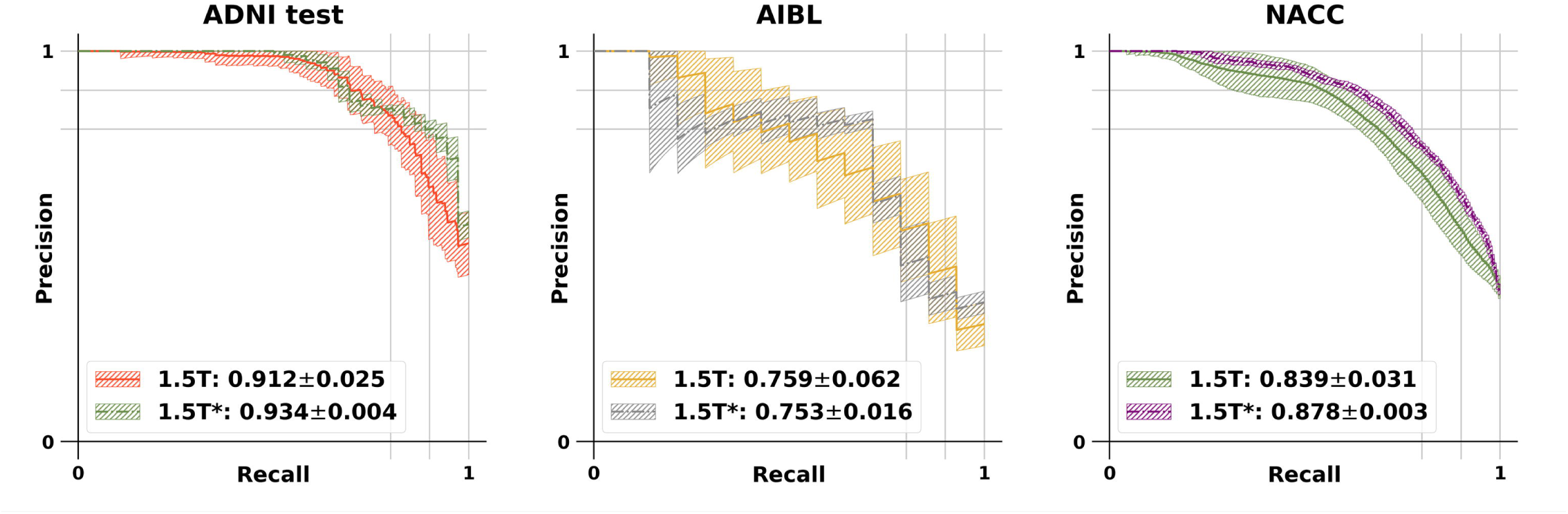
Performance of the FCN models. (a) Sensitivity-specificity (SS) and (b) precision-recall (PR) curves comparing the FCN models predicting Alzheimer’s disease status. One FCN model was developed using the 1.5T scans and the other using the 1.5T* images. Model performance is shown on all three datasets (ADNI test, AIBL and NACC).

## Discussion

Our deep learning pipeline involved training of a GAN to learn from 1.5T and 3T scans obtained from the same subjects, and an FCN to simultaneously predict AD status directly from the generated 1.5T* images from the GAN (Figure 1). Simultaneous minimization of losses from the GAN and the FCN enabled us to achieve improvements in MR image quality and AD classification performance. Moreover, volumetric patch-level training of the GAN and the FCN models turned out to be computationally efficient, where the size of the patches was the same as the receptive field of the FCN. Importantly, access to 1.5T and 3T scans on the same subjects taken at the same time was crucial to develop 1.5T* images, without the influence of potential confounding factors such as scan timing and the scanner itself. Also, both the AIBL and NACC datasets served as good, independent datasets for model validation, allowing use of the same criterion for subject selection on all these cohorts.

Our study implication is that it is possible to go back in time to generate images of enhanced quality on disease cohorts that used 1.5T scanners, and use them along with the more recent datasets developed using 3T scanners. This would allow us to reconstruct the earliest phases of AD, and build a more detailed model of disease progression than would otherwise be possible using multiple datasets. Our proposed adversarial framework can also be extended to process other medical imaging datasets and organ systems, where there is a need to normalize longitudinal scans that can conform to a single technological standard of precision, so that more accurate models of disease progression can be developed.

There is a putative link between MRI scans with high quality (defined using SNR, etc.) obtained from latest instrument-level advancements and their ability to better delineate structural aspects that manifest in various diseases. It is appealing to embrace MR images of high SNR to improve detection of structural changes in the human brain. This seemingly advantageous technological progress poses a conundrum – models created using MR images at early time points using older technology may not be sufficiently accurate in terms of predicting AD status. Further, longitudinal changes determined from 1.5T and 3T scans due to neurodegeneration cannot be confounded by increased sensitivity due to higher magnet strength. This becomes more important in the case of aging individuals who could benefit from more accurate assessment of cognitive status early in their lives. While there is not yet any available drug treatment for treating cognitive abnormalities with insidious onset such as AD, research indicates that delaying onset will cut an individual’s risk for diagnosis (15, 16). Using the GAN framework and MR images of different MFS, we developed a model to generate images of improved quality and predict AD status of individuals prior to reaching clinical disease diagnostic thresholds with greater accuracy than what would have been possible using models generated using images with single MFS alone.

Our study has a few limitations. First, the sample size used for GAN model training was small (n=151), as only a limited number of cases had both 1.5T and 3T imaging done at the same time. It is possible to generate a more robust GAN model if such data is available on a larger number of cases. Both the GAN and FCN models were designed to have specific architectures. More optimized architectures can be constructed and this could alter the performance of the models. We used well-known no-reference algorithms such as SNR, BRISQUE and NIQE to evaluate image quality on the images, and additional quality metrics could be explored. Even though the BRISQUE evaluator was not designed to evaluate medical images, we still used the BRISQUE evaluator to explore whether distortion features from natural images could statistically distinguish any subtle differences between MRIs collected with various magnetic field strengths. We observed that the image quality of both 3T scans and generated 1.5T* scans statistically outperformed that of the original 1.5T scans. Nevertheless, the ability of the GAN model to generate images of similar quality to that of the original scans was consistent across these metrics, and the enhanced AD classification performance was evident, as evaluated using independent test data.

In conclusion, our approach to produce high AD classification performance models and improve image quality using a GAN framework could transform the way MRI data is utilized in AD research. In research, longitudinal assessment is necessary to investigate causal mechanisms, but longitudinal data, especially from MRI, requires harmonization. Our modeling paradigm could empower longitudinal studies by enabling better use of MRI scans that are already collected.

## Data Availability

Python scripts are provided on the GitHub repository.

https://github.com/vkola-lab/gan2020

## Acknowledgments

This project was supported in part by the National Center for Advancing Translational Sciences, National Institutes of Health, through BU-CTSI Grant (1UL1TR001430), a Scientist Development Grant (17SDG33670323) and a Strategically Focused Research Network (SFRN) Center Grant (20SFRN35460031) from the American Heart Association, and a Hariri Research Award from the Hariri Institute for Computing and Computational Science & Engineering at Boston University, Framingham Heart Study’s National Heart, Lung and Blood Institute contract (N01-HC-25195; HHSN268201500001I) and NIH grants (R56-AG062109, R01-AG062109, AG008122, R01-AG016495, and R01-AG033040). Additional support was provided by Boston University’s Affinity Research Collaboratives program and Boston University Alzheimer’s Disease Center (P30-AG013846).

## List of abbreviations

MRI: Magnetic Resonance Imaging
GAN: Generative Adversarial Network
CNN: Convolutional Neural Network
GPU: Graphical Processing Unit
SNR: Signal to Noise Ratio
SSIM: Structural Similarity Index Metric
BRISQUE: Blind/Referenceless Image Spatial Quality Evaluator
NIQE: Natural Image Quality Evaluator

## Supplemental information

### Generative adversarial framework

The framework consists of three models, i.e., generator, discriminator and a disease classifier. The generator takes 1.5T MRI as input and produces a scan where we call enhanced 1.5T* MRI. The discriminator tries to distinguish the generated 1.5T* scan from 3T scan and sends difference information back to the generator. Thus, by performing the two-player game between generator and discriminator, generator is able to produce more and more realistic scans with the aid from discriminator and at the meanwhile, discriminator also got improved in classifying the subtle difference between the generated 1.5T* and real 3T scans. The loss functions for the discriminator is the binary cross entropy loss as shown below:

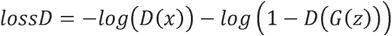

where x represents real 3T scans, z represents 1.5T scan, G and D refer to the generator and discriminator models, respectively. Note that G(z) is the generated 1.5T* scan. In this setting, the discriminator are trained by assigning 3T as real images and 1.5T* as fake images.

The generator is supposed to generate scans which discriminator might consider as real scans, thus the first part of the loss function of the generator is:

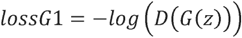

Since we trained the GAN model by sending in paired 1.5T and 3T scans from same subjects, the generated images should be ideally the same as 3T scans. Thus, the second part of generator’s loss function is L1 norm between generated 1.5T* and 3T scans:

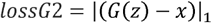

Besides these two parts, the GAN framework was trained together with an auxiliary disease classifier, which is another fully convolutional network, denoted as FCN. The FCN classifies generated 1.5T* scans according to the disease label, which is either AD, representing Alzheimer’s disease, or NC, representing normal cognition. The disease classification loss will be back-propagated to the generator as well, which contributes to the third part of the generator’s loss function:

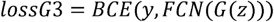

where BCE represents binary cross entropy loss and the disease ground truth y take values below:

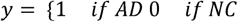

Combining all the three terms, the total loss of the generator is:

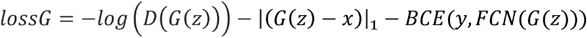

Note that while the generator and discriminator training, the FCN was also trained from scratch by learning the same BCE loss below:

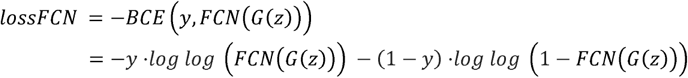

By learning from the generated scans which own large variance due to the stochastic nature of the deep learning generator, FCN can benefit from the data variety as a form of efficient GAN-based data augmentation.

### Supplemental figures and tables

**Supplement Table 1:** Specific between group differences in magnetic field strength (1.5T, 3T, 1.5T*) evaluated using Tukey’s post hoc procedure across the image quality measures SNR, BRISQUE and NIQE in the ADNI-training, testing and validation data.

| ADNI-training |  |  | ADNI-validation |  | ADNI-testing |  |
| --- | --- | --- | --- | --- | --- | --- |
| SNR |  |  |  |  |  |  |
|  | 1.5T | 3T | 1.5T | 3T | 1.5T | 3T |
| 1.5T* | <0.0001 | <0.0001 | <0.0001 | <0.0001 | <0.0001 | <0.0001 |
| 3T | 0.89 |  | 0.79 |  | 0.92 |  |
| BRISQUE |  |  |  |  |  |  |
|  | 1.5T | 3T | 1.5T | 3T | 1.5T | 3T |
| 1.5T* | <0.0001 | 0.04 | <0.0001 | 0.02 | <0.0001 | 0.09 |
| 3T | 0.0006 |  | 0.002 |  | 0.02 |  |
| NIQE |  |  |  |  |  |  |
|  | 1.5T | 3T | 1.5T | 3T | 1.5T | 3T |
| **1.5T\*** | <0.0001 | 0.35 | <0.0001 | 0.57 | <0.0001 | 0.82 |
| **3T** | <0.0001 |  | <0.0001 |  | <0.0001 |  |

In order to identify specific between group differences in magnetic field strength, we used Tukey’s post hoc procedure (Supplement Table 1). We evaluated the between group differences in the ADNI data across the training, validation and testing splits. First, we assessed whether the mean image quality of the 1.5T* and 3T scans was better than the 1.5T scans. We found that the 1.5T* group had a significantly better mean image quality (p <0.0001) than 1.5T scans measured using SNR, BRISQUE and NIQE across the ADNI training, validation and testing data. On the other hand, the mean image quality of 3T scans was only significantly better than the 1.5T scans on the BRISQUE and NIQE measures, but was not significant using the SNR measure in any of the ADNI data splits.

Next, we assessed the whether the mean image quality of the 1.5T* scans was better than the 3T scans (Supplement Table 1). We found that the mean image quality measured using SNR in the 1.5T* category was significantly better than the 3T scans across all three data splits at <0.0001 level of significance. Using the BRISQUE measure, we found that the mean image quality in the 1.5T* category was significantly better than the 3T scans at 0.05 level of significant in the ADNI training (p=0.04) and validation data (p=0.02), but not in the ADNI test data (p=0.09). However, the mean image quality in the 1.5T* category was not significantly better than the 3T scans in any of the ADNI data splits, using the NIQE image quality measure.

**Supplement Figure 1 (S1).**
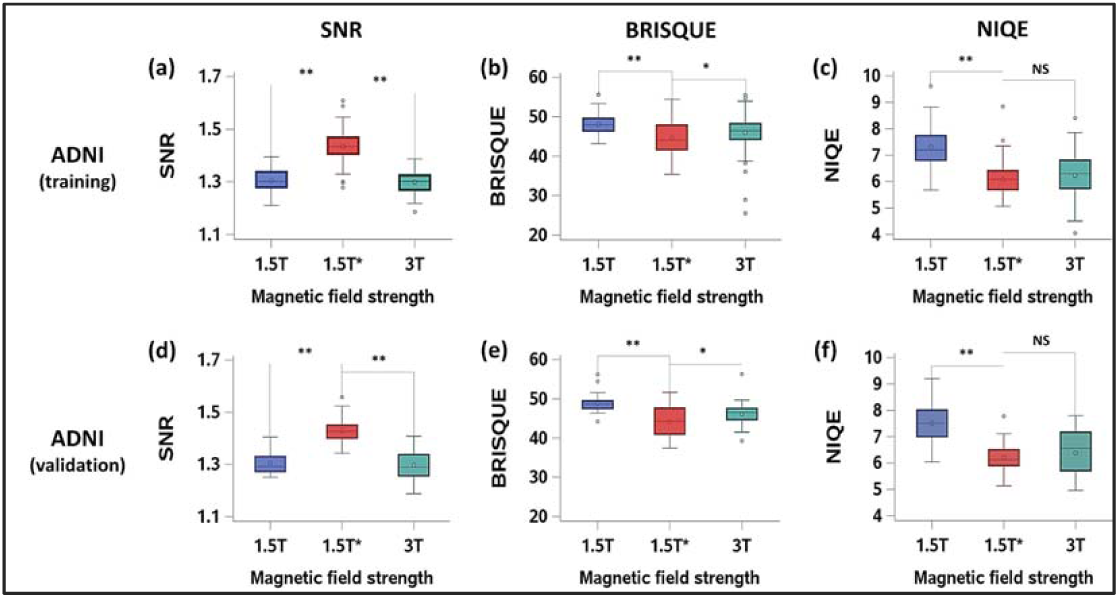
ANOVA to assess the mean image quality using SNR, BRISQUE and NIQE in the ADNI-training (a-c) and ADNI-validation (d-f) data. *: Statistical significance at p<0.05 level **: Statistical significance at p<0.0001 level

In order to determine whether mean image quality of images in the ADNI-test and ADNI-validation data differed across the different magnetic field strengths (1.5T, 3T and 1.5T*), we performed the ANOVA analysis (Figure S1). Image quality measures included SNR, BRISQUE and NIQE. We found significant evidence of an overall difference in mean image quality using all three measures between 1.5T, 3T and 1.5T* scans at 0.05 level of significance.

